# Implementing a consultation service for translating genomic research findings into the clinic: lessons from the SickKids Genome Board

**DOI:** 10.1101/2024.11.18.24317509

**Authors:** Amy Pan, Kenzie Pulsifer, Michelle Axford, Lena Dolman, Bailey Gallinger, Eriskay Liston, Elizabeth Stephenson, Anna Szuto, Laura Zahavich, Gregory Costain

## Abstract

**Objectives:** Genome-wide sequencing (GWS) is now used across the breadth of paediatric research. There is a greater potential to identify unexpected, clinically relevant findings with GWS than with the targeted genetic techniques used in prior decades. Individual research teams may not have the expertise to evaluate and manage these findings. The Hospital for Sick Children (SickKids) Genome Board is a no-cost consultation service for researchers with questions arising from genetic aspects of their studies.

**Methods:** We reviewed all submissions to and recommendations from the Genome Board over the first four years, to identify common questions, themes, and trends.

**Results:** There were 67 submissions, and a year-over-year increase in volumes. The most common request (60%) was to assess variants identified by GWS for pathogenicity, clinical actionability, and returnability to a study participant. Overall, 23 of 48 reviewed variants were recommended for clinical confirmation and return with genetic counselling. Other categories of submissions included requests to researchers from study participants to release their “raw” genomic data, and for input on protocols related to clinical translation of findings.

**Conclusion:** The Genome Board provides a generalizable model for centralized triage of clinical questions arising from genomic research at a paediatric centre. Clinicians should be aware that patient participation in genetic research studies can have downstream consequences for their healthcare.

## INTRODUCTION

Genome-wide sequencing (GWS), including both exome sequencing and genome sequencing, is now widely used in paediatric research (1–5). GWS has the potential to identify findings of possible or definite clinical relevance (1,6,7). These may be “primary” results (i.e., relevant to the phenotype under investigation), or “secondary” (6) or “incidental” (8) results (i.e., unexpected non-primary results that are intentionally or unintentionally identified, respectively). Professional societies (e.g., American College of Medical Genetics and Genomics [ACMG]) and large-scale community resources (e.g., Clinical Genome Resource [ClinGen]; Global Alliance for Genomics & Health [GA4GH]) provide guidance on research best practices (9), variant classification (10,11), disease-gene associations (11), and clinical actionability (11,12). Genetics care providers like medical geneticists, genetic counsellors, and clinical laboratory directors have expertise in interpreting and managing GWS results (7). In contrast, individual research groups may not have the up-to-date training and knowledge to safely navigate the landscape of clinical translatability (13).

At our institution (The Hospital for Sick Children; SickKids®), a significant and growing number of active research studies involve the use of GWS. The Research Ethics Board (REB) recognized the need for studies utilizing advanced genomic technologies to have an established pathway to manage potential findings. In response, the Genome Board (comprised of authors M.A., G.C., E.L., E.S., A.S.), was created in 2020 to offer a no-cost consultation service for researchers at SickKids seeking advice on issues relating to genomic research. The Genome Board aims to ensure consistency and transparency of genomic research practices, through streamlining access to recommendations regarding potential clinical relevance of specific genomic findings, and through promoting genomic research result confirmation and disclosure that meet clinical standard of care. The Board’s members have complementary expertise in clinical genetics, molecular and laboratory genetics, genetic counselling, and research ethics, with ad-hoc subject matter experts invited when needed to participate in specific case reviews.

The specific activities and overall impact of the Board have not been previously described. In this study we describe the submissions to, and resulting feedback and recommendations from, the Board over a four-year period. Given the increasing use of genetic technologies in research, the implementation of a genomic review group at the institutional level can provide a valuable service to researchers seeking support for emerging challenges and questions that arise from genomic studies.

## METHODS

This is a retrospective, single-centre cohort study approved by the REB at The Hospital for Sick Children. The Genome Board accepts any inquiries, general or specific, from researchers across the institution. Inquiries are made via an online submission form on Microsoft Power Apps. Submitting researchers include de-identified study participant information, and copies of their REB-approved protocols and consent forms, when needed. All information received from case submissions is stored in a Microsoft SharePoint list accessible only to members of the Board. The Genome Board meets monthly to review case submissions and generate feedback/recommendations to return to the submitting research team. Input from additional ad-hoc Board members with specific areas of expertise (e.g., cardiac genetics, cancer genetics) is solicited when needed. All submissions are reviewed by one member of the Board upon submission, prior to the monthly meeting. This pre-review can determine if the case may be urgent thus requiring ad-hoc full review prior to the scheduled meeting, if more information should be requested from the submitting team prior to full review, and if other expertise relevant to the request should be consulted and/or invited to attend meetings.

A summary of the Board’s feedback and recommendations is shared directly with each submitting research team and stored in the Microsoft SharePoint list. The following disclaimer is included with the written recommendations: “During the meeting, members of the Genome Board provided feedback related to queries posed by the submitting groups. This feedback was based on the information provided to them before and during the meeting. Names and other identifying data for specific research participants were purposefully NOT solicited. Genome Board members did not have access to medical records or other collateral history. Genome Board members did not have the opportunity to interview individuals about their personal and family histories, nor to perform physical examinations. For these reasons, a medical opinion could not be, and was not, provided. Genome Board members do not assume responsibility for any aspect of the research participant’s medical care. Where discussion was based around findings from a specific research study, Genome Board members did not have access to the entirety of this research data. Interpretation of genomic findings is based on current knowledge and can change over time.”

All case submissions received by the Genome Board since its inception (November 2020) to August 2024 were included in this study. Standard descriptive and summary statistics were calculated using Microsoft Excel. Qualitative data were collected using a content analysis approach to identify categories and common themes across all case submissions.

**Table 1.** Case vignettes and outcome recommendations made by the Genome Board, informed by real case submissions but with key details changed to protect patient confidentiality.

| Type of Request | Case Summary | Genome Board Recommendations |
| --- | --- | --- |
| Review of secondary or incidental variant for actionability & return | A child was recruited into a non-cardiac-focused research study that included the use of singleton (child only) exome sequencing. The consent form indicated that participants could be told about medically actionable additional findings. In the course of reviewing variants annotated as "known pathogenic" in the ClinVar database, the genome analyst identified a "likely pathogenic" variant in the *RYR2* gene associated with catecholaminergic polymorphic ventricular tachycardia (CPVT). There was no documented personal or family history of cardiac issues in the records available to the study team.<br>The Genome Board was consulted to provide guidance on whether to return this finding, and regarding next steps for the participating family. | The Genome Board recommended that this finding be returned to the family urgently. The Board agreed with the variant classification, and identified clear actionable next steps that can mediate high-risk cardiac manifestations. The Board recommended clinical confirmation of the variant and a referral to Cardiology to initiate diagnostic investigations. From a genetic counselling perspective, the Board recommended discussing generally that a variant related to heart rhythm differences was found, urging caution and against usage of the term "CPVT" to avoid causing significant anxiety in the family while the initial next steps are arranged.<br>**Impact:** An urgent referral to Cardiology to initiate an exercise stress test that will clarify this patient's risk for CPVT, where manifestations can include syncope and sudden cardiac death. Confirmation of the diagnosis and variant would enable cascade testing of family members also at risk, including the proband's parents. |
| Participant request for genomic data access | A child with a neurodevelopmental disorder underwent research-based trio (child and biological parents) genome sequencing. The study team had completed their analyses and not identified diagnostic/potentially returnable variants. The child did not have capacity to consent at the time of recruitment (the parents provided consent on the child's behalf), and it was unclear to the study team if the child would ever gain capacity in the future. The parents requested that the research team send "all" the genome sequencing data to both their new care provider and to a research group, both located in another country.<br>The Genome Board was consulted to provide guidance on if and how to release these data. | The Genome Board advised that participants have the right to their own genomic data and thus can request these data. Prior to sharing of data, consent is needed from all involved individuals and/or substitute decision makers (SDMs). In this case, the study team was advised to obtain new consent for both parents and for the child. Template documents for consent forms for data release requests are available at SickKids for study teams. The Board also advised that genomic data sharing should ideally occur through secure file transfer from SickKids directly to the requesting clinician or researcher.<br>**Impact:** A secure release of genomic data to other clinicians and researchers enables further investigations that may benefit the patient. Informed re-consent of all individuals regarding the release of data specifically is important to uphold autonomy, as consenting to participate in a genetic research study does not assume consent to share one's genetic information elsewhere, even if for the purposes of care. |
| Review of variant related to primary diagnosis | A youth with a complex and unexplained medical issue underwent research based genome wide sequencing, as one component of a study focused in discovering new causes of this medical issue. The sequencing identified a rare DNA variant, in a gene ("A") that has not yet been definitively associated with human disease but that is related to a different gene ("B") associated with these medical issues.<br>The Genome Board was consulted to provide guidance on the potential for this variant to be a primary diagnostic finding for the participant, and whether to return the result to the participant. | The Genome Board reviewed the available literature for gene A and classified it as a 'candidate gene,' defined as a gene with emerging evidence for an association with a disease phenotype that has not been formally established by curation resources (e.g. ClinGen). Thus, the Board did NOT recommend presenting the variant to the family as a diagnosis. Further exploration of this variant and gene in a research setting (e.g. functional assays, gene-matchmaker services) may generate additional evidence that helps to confirm or refute a gene-disease association.<br>**Impact:** Informed recommendations against returning a result without established disease association ensures that only research findings with the highest likelihood for clinical relevance are returned to families and able to potentially impact clinical care. |

## RESULTS

### Genome Board submissions reflected a breadth of issues pertaining to genomic research

From November 1, 2020, to August 1, 2024, the Genome Board received and reviewed 67 submissions. There was a year-over-year increase in volumes (Figure 1a). Submissions were made by 17 different individuals across 13 different research groups. The research groups were studying conditions including but not limited to developmental, neurological, rheumatological, renal, and cardiac disorders. One large-scale autism genomics research study made a majority of the submissions (n=38, 57%) (4).

**Figure 1.**
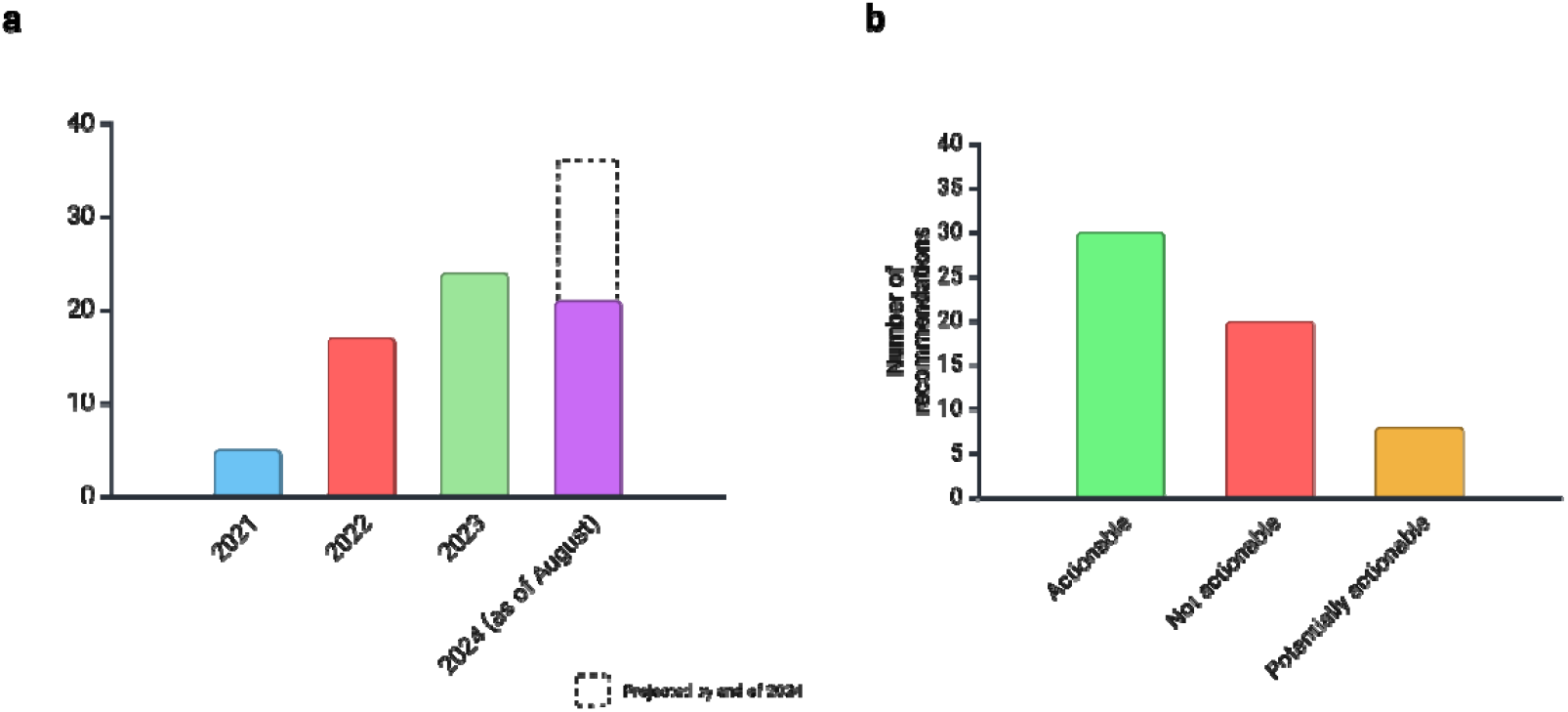
Timing and outcomes of submissions received by the SickKids Genome Board. Created with BioRender.com. (a) Number of submissions received by the Genome Board per year since its inception. No submissions were received in November 2020 or December 2020. For this report, only submissions until August 1, 2024, were included. (b) Outcome recommendations for submissions that were determined to have potential for actionability. See text for detail.

There were five main categories of submissions: (i) review of potential secondary or incidental finding for clinical actionability and returnability (n=40 submissions, 60%; 43 different variants), (ii) review of study participant access request for “raw” genomic data (n=9, 13%; including n=1 hypothetical and n=8 actual requests) (14), (3) review of potential primary finding for clinical actionability and returnability (n=8, 12%; 8 different variants), (4) query about general genomic research best practices (n=6, 9%), and (5) review of proposed genomic study design (n=4, 6%). Box 1 includes three representative case vignettes.

### Recommendations regarding returnability of DNA variants were case-dependent

Variants in 47 different genes (submitted as either a primary, secondary, or incidental finding) were reviewed by the Board. The most common phenotypic category of genes submitted for review was cardiovascular (n=12, 26%; e.g., cardiomyopathy, arrythmia). Multiple variants were submitted for four genes (*BRCA2, CLCN1, MFN2, RNF213*). Nine genes (19%) were on the ACMG Secondary Findings v3.2 gene list (12), and 17 genes (36%) had been assessed for paediatric and/or adult actionability by ClinGen (11). The Genome Board recommended returning 23 (48%) findings to participants/families, conditionally returning 8 (17%) findings (e.g., return if patient not previously assessed for condition X or not previously seen by specialist Y), and against returning 20 (42%) findings.

The Board considered the following lines of evidence in making recommendations for return of the 23 findings: (i) variant classification (e.g., pathogenic or likely pathogenic), (ii) medical actionability (e.g., clear guidelines for management and/or screening for individuals at-risk for, or diagnosed with, the associated condition), and (iii) implications for family members (e.g., initiation of cascade testing or screening for relatives). Reasons not to return findings included: (i) unclear disease association (e.g., weak or unclear gene-or variant-disease association, including candidate genes), (ii) unclear or lack of medical actionability (e.g., no guidelines or evidence for management and/or screening, including evidence indicating that management would not change), (iii) uncertain variant classification (conflicting or insufficient evidence for variant pathogenicity) (15), and (iv) poor phenotype fit for variant to be a primary diagnostic finding.

### Other considerations raised in the context of paediatric genomic research

An additional 8 (14%) of the initial submissions had potential for direct participant impact (“actionability”) in the form of requests for data release. The Genome Board recommended releasing data to participants for 7 (88%) and against releasing data for 1 (12%), as the family requested access to information that was not permitted in accordance with data governance policies. A one-page consent form template for return of genomic data is available from our institution upon request.

With respect to submissions relating to genomic research best practices and study design, feedback from the Genome Board included the sharing of existing resources (e.g., from ACMG, ClinGen, REB, institutional legal services) and recommendations to include / exclude certain elements in protocol designs (e.g., specific language use in consent forms (16)).

## DISCUSSION

The Genome Board is a genomics consultation service developed and implemented at SickKids in late 2020. Over a four-year period, the Genome Board received and reviewed 67 submissions from a diverse group of intra-institutional research teams seeking guidance on issues related to genomic research. Genomic research can and often does generate findings that warrant downstream clinical attention. Resolution of queries and formulation of feedback/ recommendations required consideration of principles including but not limited to: variant classification and gene-disease association, genotype-phenotype correlation, medical actionability, and research and clinical ethics considerations (e.g., rights to owning one’s data, patient autonomy and informed consent, protection of patient privacy).

### The Genome Board offers a valuable and increasingly utilized service to institutional research groups by guiding the resolution of challenges associated with genomic research

Since its inception, the Genome Board has been utilized increasingly across multiple disciplines within the institution. The gradual implementation of genomic components into more research protocols results in more genomic-related queries and concerns that may fall out of the scope of the organizing team. Targeted promotional efforts within the institution has increased visibility and awareness of the Genome Board, and repeated submissions from the same submitting team may indicate satisfaction with and trust in the service offered. These results demonstrate a growing interest in and need for a genomics consultation service, and we suspect a similar demand at other paediatric academic institutions. In this study, most submitting research teams did not have the volume of questions to justify significant investment in their own study-specific genomics review group. The availability and accessibility of genomics expertise on an institutional level may empower research teams to incorporate genomic components into their study designs.

The Genome Board has specifically provided guidance on many research findings of potential actionability. Secondary findings are a recurrent notable potential benefit — or burden — derived from GWS (17,18). The current clinical standard of the ACMG is to report known or predicted disease-causing variants in genes that cause medically actionable and highly penetrant conditions. Guidelines, however, cannot assess whether specific additional findings (either “secondary” or “incidental”) discovered by a research group meet the threshold for returnability to families and, if so, the appropriate next steps for the variant or counselling approaches for result disclosure. Furthermore, the advent of technology and migration of health information onto electronic-based medical systems may lead to greater desire for and capacity to access one’s own healthcare data. Genomic data, in particular because of its inherent identifiability, warrants greater caution to protect privacy. Developing consistent policies for the release of genomic data, including the establishment of appropriate informed consent procedures, will be an important focus for research groups moving forward as the number of such requests from participants increases. The submissions to the Genome Board highlight the importance of developing local, provincial/territorial, and/or national guidelines to address recurrent issues related to genomic research, to ensure consistency and transparency in practice.

Our study has several limitations. The submissions received at SickKids may not be representative of the questions and challenges at other institutions. Questions about the amenability of DNA findings to pharmacogenetics (19) or genetic therapy development (20,21) are expected to increase in the coming years. The outcomes of cases following the return of recommendations were unknown. Specifically, we were unable to confirm if the Genome Board’s recommendations were followed by the study team, the level of submitter satisfaction with the service provided, or if additional informative patient information was discovered during or after the review process. The Genome Board’s recommendations with respect to a specific variant or query may change over time. However, one author (A.P.) re-reviewed all submissions during the drafting of this manuscript, and there were none where the variant interpretation and/or rationale for/against a specific recommendation had changed enough to warrant re-review by the Board.

In summary, implementing a consultation service for translating genomic research findings into the clinic at a large pediatric academic centre was necessary, feasible, and required minimal resources beyond expert volunteers. The Genome Board is one model for adjudicating and providing guidance for queries relating to genomic research, through leveraging expertise from clinical areas. Improving the quality of institutional genomic research practice and enabling findings to appropriately impact care outcomes can benefit researchers, care teams, and participants and their families. Precision Child Health (22) and other large-scale genomics-related initiatives in paediatrics will necessitate enhanced coordination between clinical and research services, particularly as genomic data is increasingly sought after, generated, and validated for use in care.

## Data Availability

All data produced in the present study are available upon reasonable request to the authors.

## Acknowledgements and funding

We thank the research teams at SickKids for submitting questions to the Genome Board, and ad hoc Genome Board members for their input on specific submissions. Funding for genetic counselling and administrative support was provided by Novartis

## Notes

### Competing Interest Statement

The authors have declared no competing interest.

### Funding Statement

This study was funded by Novartis to support genetic counselling and administrative support.

### Author Declarations

The Hospital for Sick Children (SickKids) Research Ethics Board gave ethical approval for this work.

